# Sense and Sensibility: Sensible Quality Control and Data Decision-Making in Passive Sensing Data for Adolescent Substance Use Risk Prediction

**DOI:** 10.64898/2026.10.06.26364884

**Authors:** Anni Hong, Jihui L. Diaz, Traci M. Kennedy, Frances L. Wang

## Abstract

**Background:** Passively collected smartphone and wearable data, combined with genomic information, may enable scalable early identification of youth who are at risk for substance use disorder. However, heterogeneous data collected in naturalistic settings limit researcher control over data quality, requiring substantial preprocessing. Data cleaning and feature engineering decisions can meaningfully influence analytic outcomes yet are often minimally described or omitted in published work.

**Objectives:** We provide rationales and recommendations for common preprocessing challenges in passive sensing research, including establishing inclusion criteria when “outliers” may reflect true behavioral variability, analyzing differences across smartphone operating systems, handling observations collected outside protocol windows, and engineering features for machine learning applications.

**Methods:** We illustrate principled approaches to these challenges using smartphone and Fitbit data from the Adolescent Brain Cognitive Development^SM^ Study (ABCD). The sample was drawn from the available half of the year-4 follow-up (data release 5.1; N = 4,754; ages 13–14).

**Results:** Extreme outlying observations were concentrated among low-engagement participants with few observed days. Smartphone usage among Apple users appeared under-recorded relative to Android users when compared with self-report measures. Fitbit-derived behavior differed systematically between observations collected within versus outside protocol windows. We also identified potential errors in the study’s default preprocessing pipeline.

**Conclusion:** We recommend prioritizing person-level inclusion criteria that preserve meaningful behavioral variability, examining temporal distributions rather than assuming observations are exchangeable, and thoroughly investigating and reporting data quality issues. Although illustrated using the ABCD Study, these principles broadly apply to research using smartphones, wearables, and other digital data sources.

## Introduction

Passively sensed data from smartphones and wearable devices offer unprecedented opportunities to characterize behavior in naturalistic settings, with growing applications in physical and mental health research[1, 2]. Large-scale studies such as the Adolescent Brain Cognitive Development (ABCD) Study have begun incorporating these digital measures [3], creating rich multimodal datasets that capture behavior at scale.

Adolescents who initiate substance use before the age of 15 face heightened risk for substance use disorder (SUD) [4–6], underscoring the need for scalable tools to identify at-risk individuals early. Our goal was to analyze passively collected sleep, physical activity, and smartphone usage data alongside polygenic risk scores using machine learning to identify “genetic screenotypes”—clusters of adolescents with distinct digital and genomic profiles that may inform substance use risk prediction. However, the promise of passive sensing data comes with substantial preprocessing challenges [7] that are often underreported and underexplored in literature. While many studies using passive sensing data report final sample sizes and basic inclusion criteria, the decision-making processes and rationales underlying data cleaning and feature engineering are rarely discussed in detail.

For instance, an important aspect of preprocessing is deciding how to treat outliers or extreme data points that fall into unlikely ranges and establishing inclusion criteria to ensure data quality. Typically, studies using passively sensed data employ a variable-based approach to removing observations based on predetermined cutoffs, applying exclusion criteria such as different wear time cutoffs [8], sleep durations <3 hours and >15 hours [9], short sleep duration 3 standard deviations below the group mean, or Fitbit steps <1000 [10]. However, it is not clear in what cases these outlying observations are erroneous versus accurate data reflecting aberrant sleep and activity patterns. Since individuals with such irregular sleep and activity patterns may be at higher risk for mental health issues and substance initiation [11–13], it may be important to consider potentially extreme observations. For example, mixed “typical” and “extreme” sleep and physical activity patterns within an individual may reflect potentially prognostic irregularity; in this case, variable-based cut-offs may artificially inflate sleep quality measures and bias downstream analyses. Moreover, establishing objective ranges is challenging: what constitutes reasonable bedtimes for adolescents? Should thresholds differ between weekdays and weekends? Given these limitations of variable-based approaches, we examined patterns of extreme values in our own data to inform our data quality strategy.

Managing data heterogeneities due to differences in operating systems (OS) is another well-documented challenge in working with passively sensed smartphone data [14–16]. For example, due to privacy restrictions on Apple’s iOS devices, it is known that the system used to collect smartphone data were unable to capture keyboard usage on the iOS native keyboards [17, 18], thus requiring iOS users to download the system-specific keyboard. Such constraint affects ABCD[19, 20] and other studies [21, 22] relying on the same collection system. The system-specific keyboard differed from the native iOS keyboard and may have been challenging or frustrating for users to adapt to and employ; therefore, it is suspected that Apple users at times disabled the non-native keyboard required for keystroke data collection in iOS devices in favor of the iOS keyboard [19]. This discrepancy in data collection might lead to inflation of low usage days for Apple users. For example, teens who use their phones frequently might still only have a few keystroke recordings. Thus, we examined whether self-reported data on smartphone usage corresponded with keyboard usage and whether such associations differed by OS.

Passively sensed data collected over time poses another source of data quality issues. Data collection could start before, or continue after, the official protocol/study window, resulting in varying amounts of out-of-protocol data across participants. This additional data may provide useful information on the participants’ engagement profiles [23, 24] and increase the effective sample size by including additional observation days. However, individuals may behave differently outside of the study window with less researcher monitoring and greater risk of disengagement or fatigue. Researchers may also receive aggregated data containing all observation days, regardless of protocol period, as we encountered in the ABCD smartphone summary dataset. Because the Fitbit and smartphone data were collected from the same individuals over the same period, we compared the Fitbit data from protocol versus non-protocol periods as a proxy for the possible limitations of the aggregated smartphone dataset, and to inform our decision about including non-protocol Fitbit data.

Facing unique challenges working with passive sensing data, researchers often need to make ad-hoc preprocessing decisions due to the lack of consensus in the literature. Thus, providing detailed documentation on such decisions and their rationales are crucial to reproducibility and interpretability. In this paper, we illustrate our decision-making process in detail through an example research project using smartphone and Fitbit data from the ABCD Study. Specifically, in order to prepare these data for clustering, we confronted several key decision points: (1) establishing data inclusion criteria when “outliers” may reflect true behavioral variation rather than measurement error, (2) dealing with discrepancies in data collection due to difference in operating systems of smartphones, (3) handling temporal heterogeneity including observations collected outside designated protocol periods, and (4) engineering features appropriate for machine learning applications. By providing detailed discussions and transparently documenting our assumptions, our work not only offers practical guidance for researchers using the ABCD dataset but also serves as a case study for others working with similar large-scale passive sensing datasets.

## Method

### Data

The sample for this study was collected as a part of the **Adolescent Brain Cognitive Development^SM^ Study (ABCD Study^®^)** (https://abcdstudy.org/), a large, publicly available dataset of youth (*N*=11,875 at baseline; 9–10 years old) accompanied by an online data dictionary, documentation, and data processing notes. The ABCD study is a longitudinal study spanning 21 research sites from 17 states, tracking the biological and behavioral development of participants to adulthood [25]. Specifically, we focused on data drawn from the available half of the year 4 follow-up (data release 5.1[26], *N*=4,754; ages 13-14). Participants were included if they participated in additional passive sensing studies with passively collected smartphone (if they had compatible devices [19]) and Fitbit sleep and physical activity data (Fitbit devices provided), along with substance use and demographic/survey information required for the project, on at least one weekend and one weekday (N = 904). The preprocessing steps detailed in the Method section were conducted on the smartphone and Fitbit datasets for these 904 participants. The average age of the preprocessing sample of participants were 14.1 years (SD: 0.7), 52% were female, and the race/ethnicity breakdown is as follows: participants identified as: White: 524 (58.0%), Non-white Hispanic: 177 (19.6%), Black: 89 (9.8%), Asian: 17 (1.9%), Other: 97 (10.7%).

**Figure 1.**
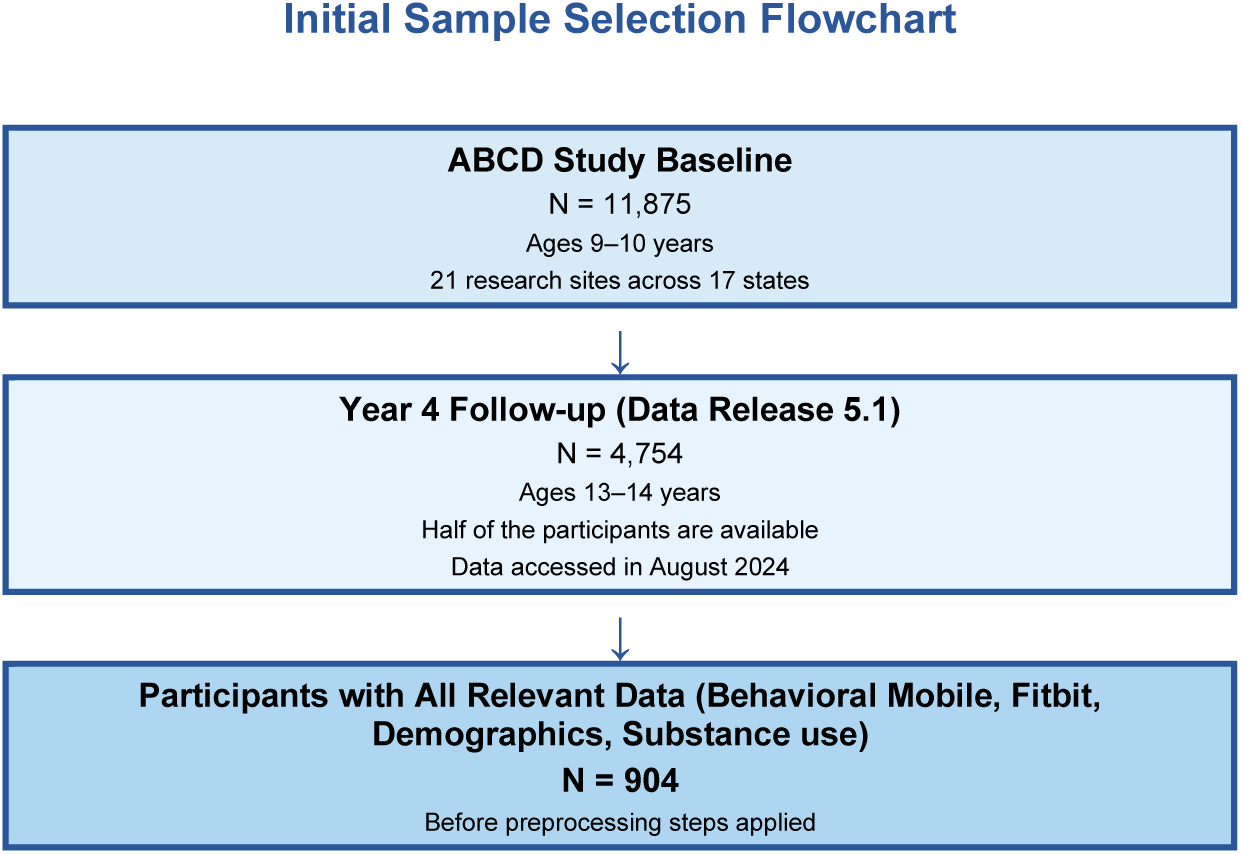
Initial Sample Selection Flowchart. ABCD = Adolescent Brain and Cognitive Development Study. Sample reflects available data at time of study (August 2024).

### Measures

#### Smartphone data (EARS)

For three weeks, an ABCD-specific version of the Effortless Assessment Research System (EARS) [17, 18] ran on participants’ smartphones. It obtained keystroke data from both Android (via native Android keyboard) and iPhone (via EARS keyboard) users.

The raw passive sensing data was previously preprocessed by Alexander et al[19] and the summary datasets were made available for public use. We used these data, which was based on the following daily keystroke measures: *key:* daily count of keystrokes; *ses*: number of sessions (a session is defined as a sequence of keystrokes with the same application no more than 60 seconds apart), and *min:* the total number of minutes spent in all sessions in a day, see Alexander et al[19] for full details.

#### Sleep and Physical Activity (Fitbit)

ABCD participants wore the Fitbit Charge HR for 3 consecutive weeks throughout the day and night. Fitbit measured a variety of physical activity and sleep-related variables calculated based on a proprietary algorithm[27]. Fitbit captured physical activity measures including total steps and time spent in active levels calculated based on heart rate (e.g., sedentary, lightly active, fairly active, and very active minutes). The sleep measures included: timing variables (going to bed, falling asleep, waking up, and getting up), sleep duration, and sleep stages (i.e., number of minutes spent in light, deep, REM, and awake). We used the passively sensed Fitbit data collected in 2021 (year 4 follow-up) summarized to the day level by the ABCD team. Note that while sleep and physical activities were captured through the same Fitbit device, they were preprocessed separately [28, 29].

#### Summary of Passively Sensed Digital Measures

**Table 1.**
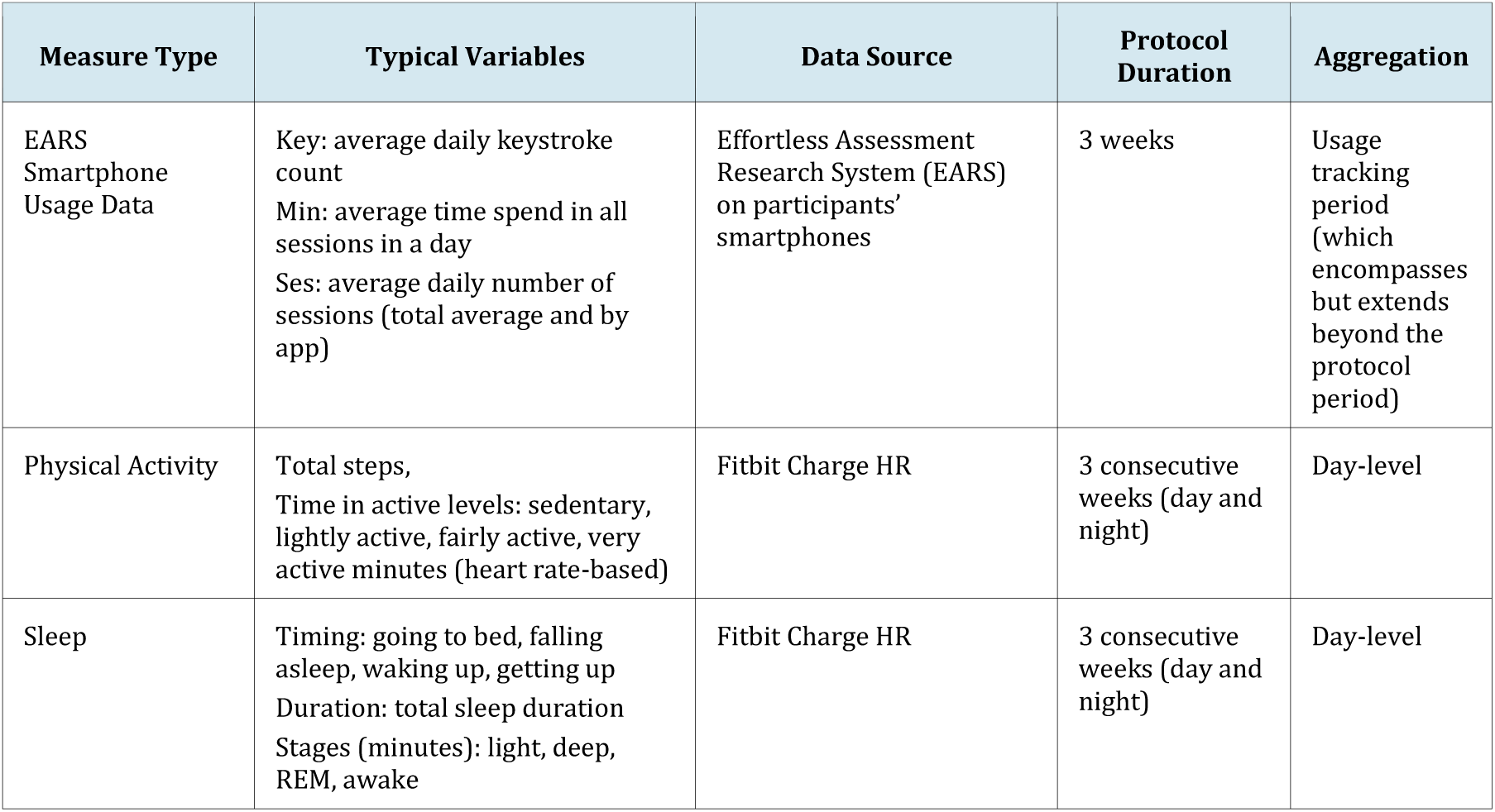
Summary of passively sensed digital measures. Fitbit sleep and physical activity data captured by same device but processed separately. All variables calculated using proprietary algorithms provided by respective device manufacturers.

### Software and Computation Environment

All preprocessing and data quality procedures were implemented in R (version 4.3.3). Core data manipulation was conducted using packages from the tidyverse ecosystem such as dplyr. Data visualization and quality assessment summaries were generated using ggplot2. R packages hms and lubridate were used to handle date/time variables and R package circular was used to compute circular statistics for timing variables in polar coordinates (see later sections for detail). Scripts used for data preprocessing are publicly available (will be made available upon publication).Results

### Outliers and Inclusion Criteria

Our first task was to establish inclusion criteria for our project to ensure data quality. In the Fitbit datasets, we identified many unlikely average daily observation values. For example, as shown in Figure 2, while the median values are reasonable, some observations of sedentary time were extreme – 0 minutes on one end of the spectrum, and near 1440 minutes (24 hours) on the other. These values seemed highly implausible given the ABCD definition of sedentary time (total daily minutes spent in activities below 1.5 metabolic equivalents [METs], as measured by Fitbit). For sleep onset times, outliers included average times between 4:00–8:00 am and 4:00–8:00 pm, representing highly atypical sleep patterns. While it might be justified to universally exclude observations with extreme values such as these, it was difficult to definitively establish acceptable ranges for sleep or activity measures that would support excluding data as erroneous, especially given the likelihood that extreme, but real, data represent important behavioral variability for capturing risk for substance initiation. Therefore, we delved deeper into the data to help guide inclusion decisions for outlying observations.

**Figure 2.**
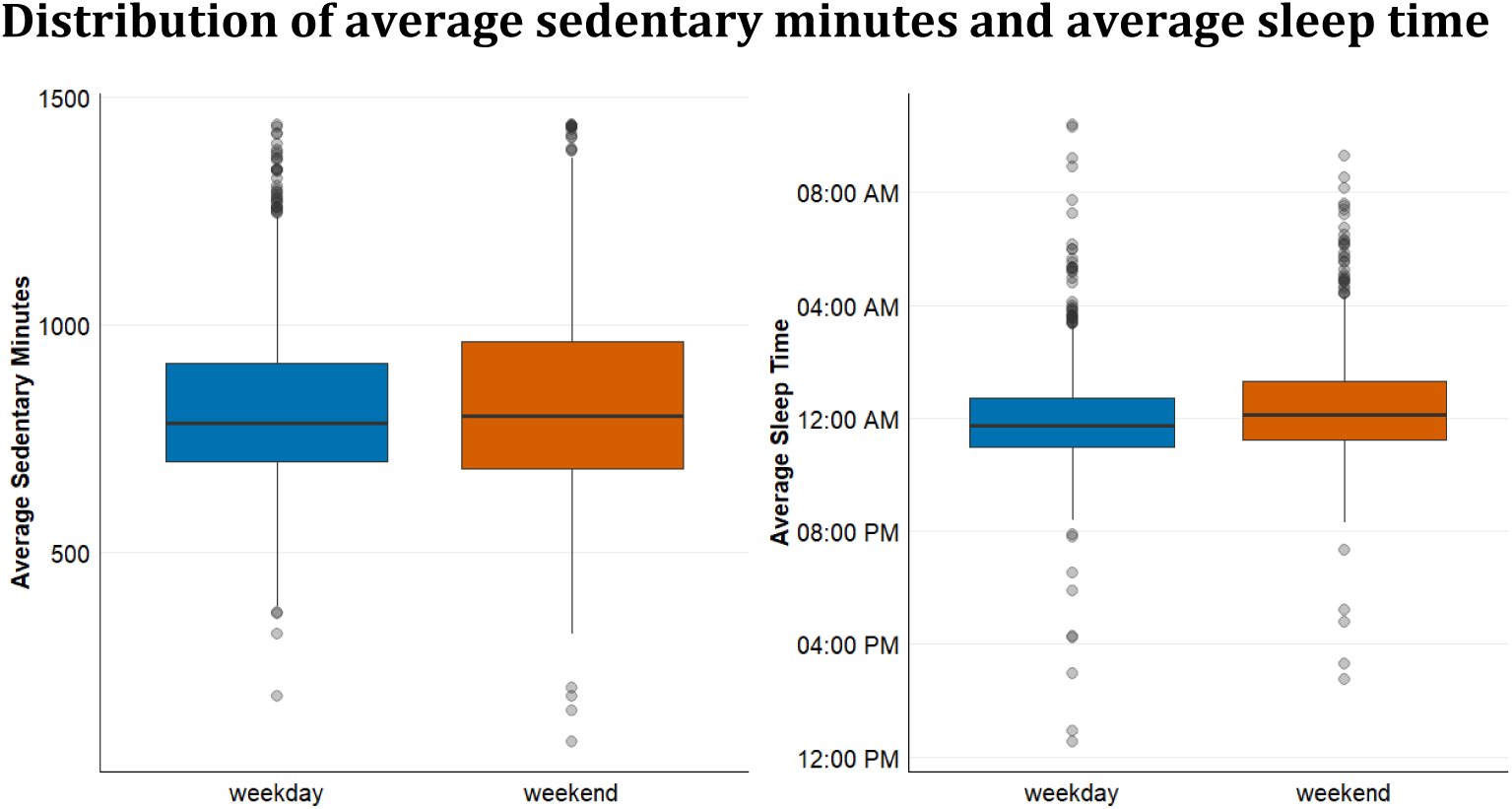
Average daily sedentary minutes (left) and sleep time (right) by weekend/weekday. Median values are reasonable; however, outliers are evident (near 24 hours and near 0 minutes for sedentary time or unusual average sleep times).

### Distribution of average sedentary minutes and average sleep time

#### Engagement-level distribution of outliers vs. all participants

To gain insight into the credibility of extreme values, we examined participant engagement as a proxy for data validity. In our dataset, we observed each participant for up to 15 weekdays and 6 weekend days out of the 21-day protocol period, though engagement varied considerably across participants, with some having substantially fewer observed days. To understand the relationship between engagement and outlying datapoints, we plotted the participant-level averages for all the Fitbit variables with the outliers (values beyond 1.5 times the interquartile range, defined as the difference between the 75th and 25th percentiles) colored based on the number of observations included in the averages (e.g., red indicates less than 5 weekdays and exactly 1 weekend day). Figure 3 illustrated that sleep and sedentary outliers tended to be red or orange indicating that extreme sleep patterns and sedentary minutes (∼24 hours or near 0 hour) are almost all from individuals with sparse observations. This pattern suggests that these values were concentrated among participants with fewer data points observed. The distribution of outliers by the engagement category (Table 2) further supports that outliers are concentrated in the low-engagement category. For every variable, the share of outliers observed for few days (Low, highlighted) far exceeds that category’s share across all participants, while high-engagement participants are nearly absent among outliers.

**Figure 3.**
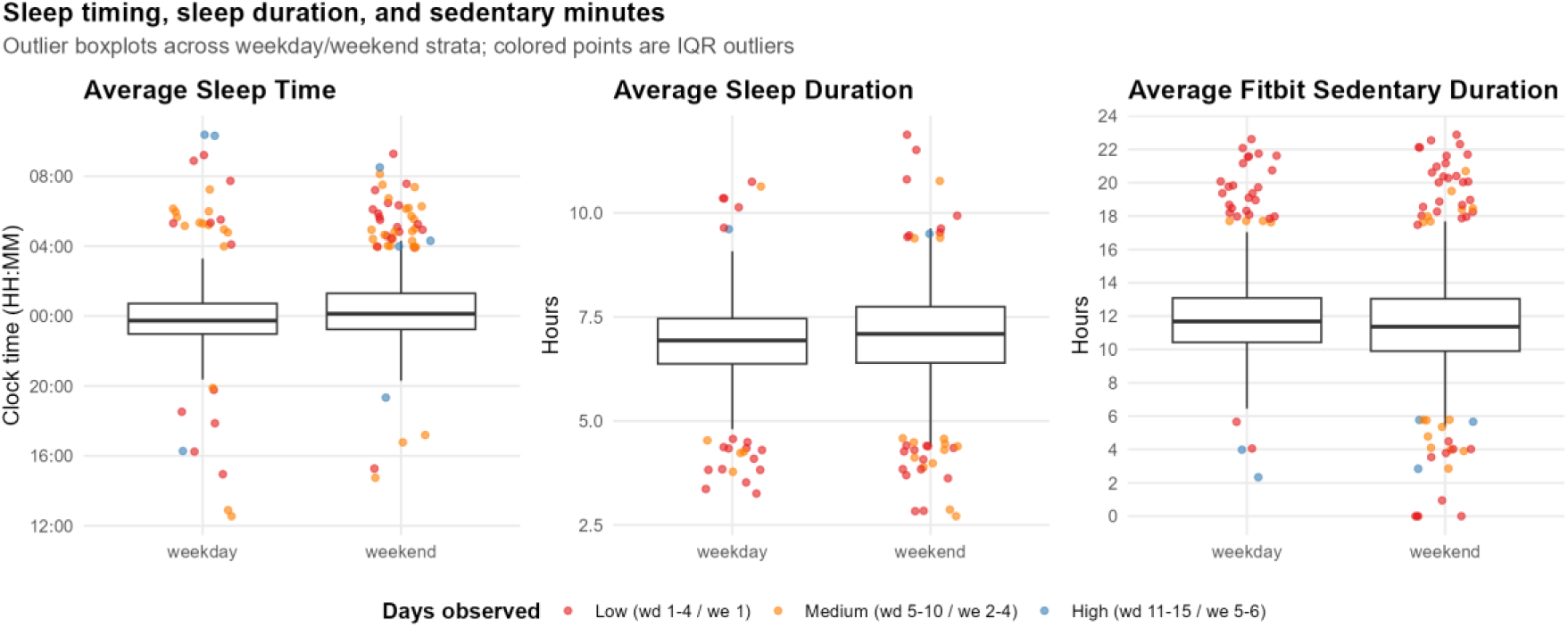
Visualizing outliers in observed average sleep time (left), sleep duration (middle), and sedentary duration (right) by number of days observed for both weekday and weekend. Red indicates a low number of observed days for a particular participant, orange for medium, blue for consistent observation over the protocol period

**Table 2.**
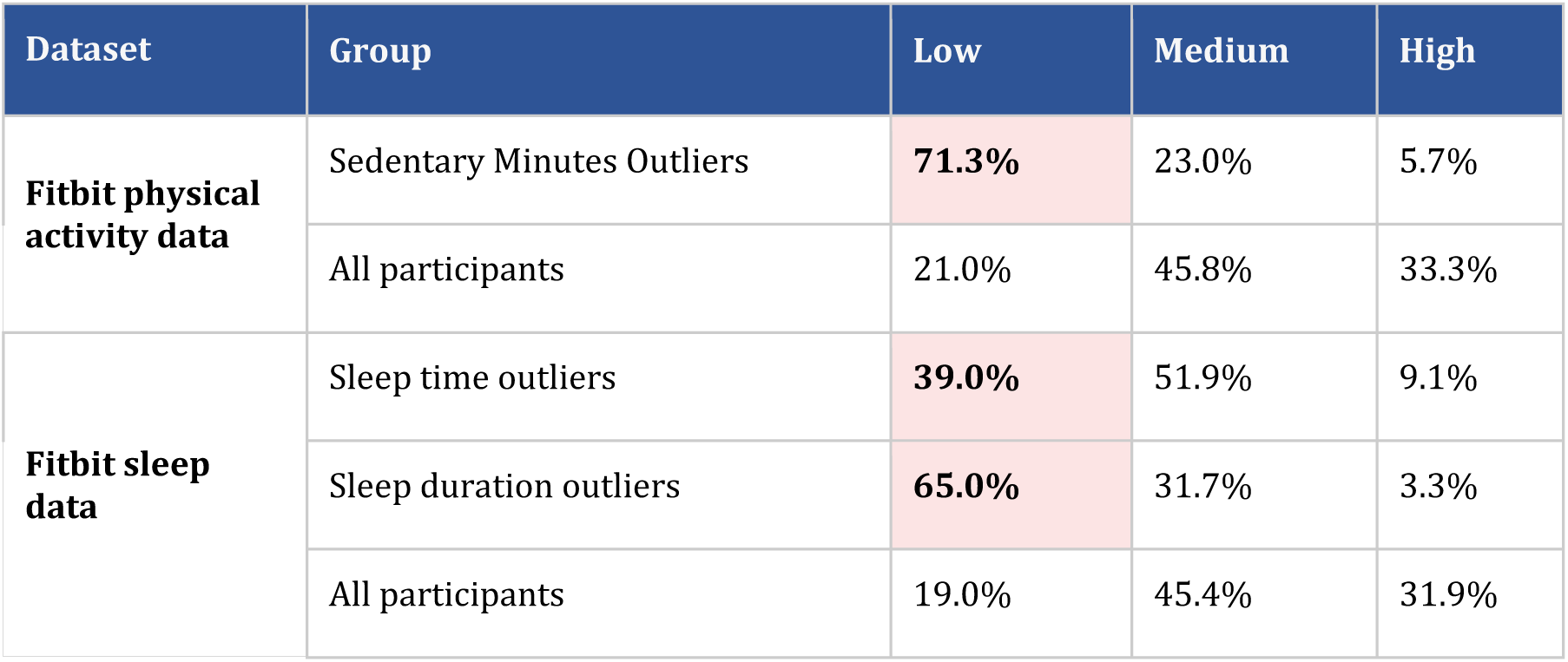
Engagement-level of outliers vs all participants. Engagement is measured by the number of days a participant was observed (Low / Medium / High).

| Dataset | Group | Low | Medium | High |
| --- | --- | --- | --- | --- |
| Fitbit physical activity data | Sedentary Minutes Outliers | 71.3% | 23.0% | 5.7% |
|  | All participants | 21.0% | 45.8% | 33.3% |
| Fitbit sleep data | Sleep time outliers | 39.0% | 51.9% | 9.1% |
|  | Sleep duration outliers | 65.0% | 31.7% | 3.3% |
|  | All participants | 19.0% | 45.4% | 31.9% |

Thus, we adopted person-based criteria including only participants with reliable data coverage, or a certain number of observations for the weekdays and weekends, rather than a variable-based approach to data inclusion. The person-based approach requires a single cutoff for the minimum number of days needed across all variables. Importantly, data points capturing poor sleep are retained if individuals meet the threshold for amount of available data across weekdays and weekends. Figure 4 illustrates how the person-based approach preserves important irregularities in the data. For example, if someone sleeps poorly most nights but occasionally gets adequate sleep—perhaps through catch-up sleep—variable-based cutoffs might remove only the extreme poor-sleep observations, misclassifying this individual as a consistently healthy sleeper. In contrast, the person-based approach would retain data indicating poor periods of sleep for individuals who provide enough data points.

**Figure 4.**
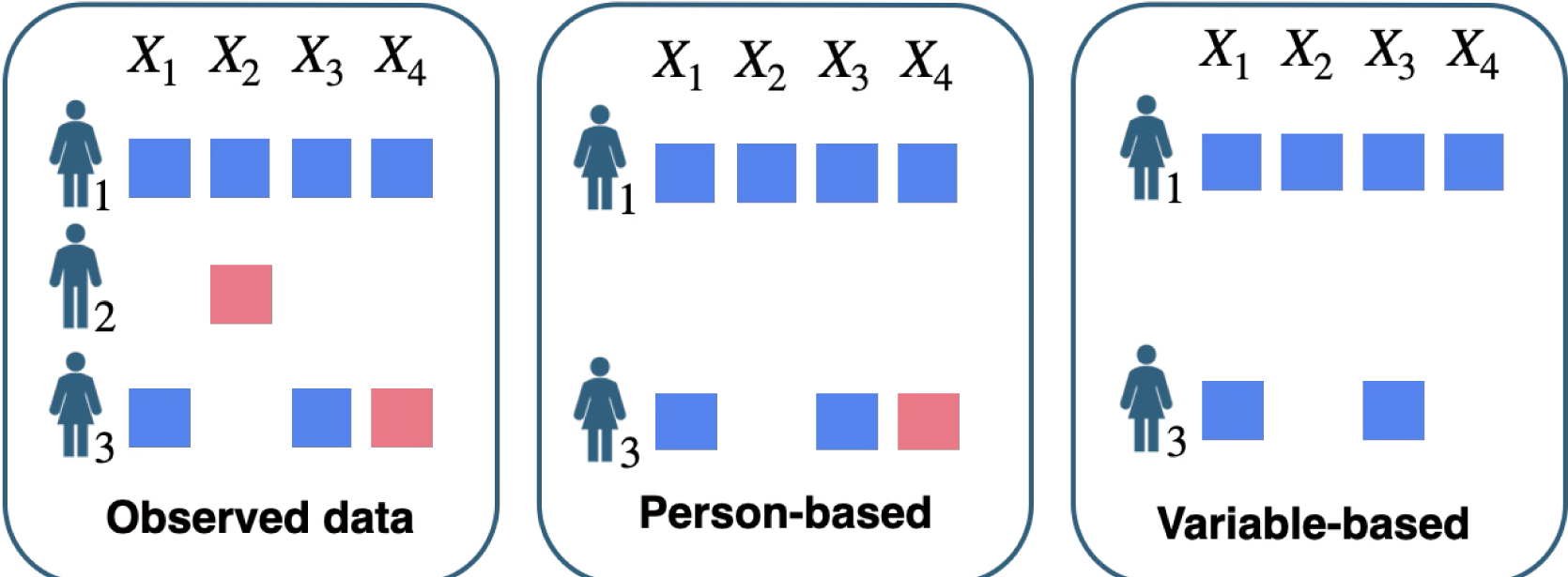
Presumed reliable data (blue), questionable quality data (pink). For individuals with mixed “typical” and “extreme” sleep patterns, variable based cut-offs may artificially inflate sleep quality measures and bias downstream analyses.

#### Illustration of person-based vs variable-based cut-offs

To determine the exact threshold for the person-based cutoff, we considered several approaches. First, we considered using the cutoff established previously by the ABCD team, who recommend only including weeks in which there were at least 4 days of data, including at least 1 weekend day [30]. However, our interest was not in week-level indices of sleep and physical activity as in other studies, but rather in treating the 3-week Fitbit period as a sampling of the participant’s activity to use as a proxy for their sleep and physical activity metrics. Therefore, we decided to only include a participant if we observed at least 5 weekdays and 2 weekend days across the 3-week Fitbit period, similar to prior studies requiring 7 nocturnal sleep events with both weekend and weekdays [31]. Moreover, combining 5 weekdays and 2 weekend days mimics a full week of data to provide a reasonable sampling of adolescents’ sleep and physical activity. Finally, measures such as social jetlag need 7 days containing both weekend days and weekdays to reliably estimate (Hasler, personal communication;[32])

### Discrepancies in Smartphone Use Data Collection Due to Differences in Mobile Device Operating Systems

In the EARS summary dataset, each participant has entirely missing data for all app category variables corresponding to their non-used operating system because Android and iOS applications were categorized differently and coded as separate variables in the dataset. To address the incompatibility of app usage data, we harmonized the categories (e.g., social, entertainment, education) between the two OSs, based on the process derived in a prior study[19]. While straightforward, this necessary data processing step required consultation with the ABCD literature to ensure accurate app categorization across OS and consistency between findings.

Another consideration was whether and how OS may have affected differences in keystroke data. We found that teens who use iPhones have been recorded with lower average keystrokes and total minutes of smartphone usage compared to Android users (Figure 5). However, as noted above, iOS users in the ABCD study were required to download the EARS keyboard to capture keystroke data. To evaluate whether the inability to use the native iOS keyboard hindered compliance with the EARS system among Apple users, we examined the relationship between keyboard usage and self-reported usage. Indeed, we found differences in this relationship across the two operating systems. As shown in Figure 6, at the lower end of usage (< 50 minutes per day), iPhone users self-reported higher smartphone usage than Android users, indicating that these iPhone users may have actually used more than what was passively recorded, assuming the propensity for self-report does not change by OS. However, it remains unclear whether this pattern is due to passive screen usage not being captured by the keyboards (i.e., passive scrolling) or if Apple users disabled the EARS keyboard, resulting in gaps in the data. Such structural differences could create systematic bias between operating systems. Ideally, this should be addressed during data collection by improving the EARS keyboard experience for iOS users or tracking when users switch away from EARS. When this is not feasible, we recommend analyzing iOS and Android data separately if sample sizes permit.

**Figure 5.**
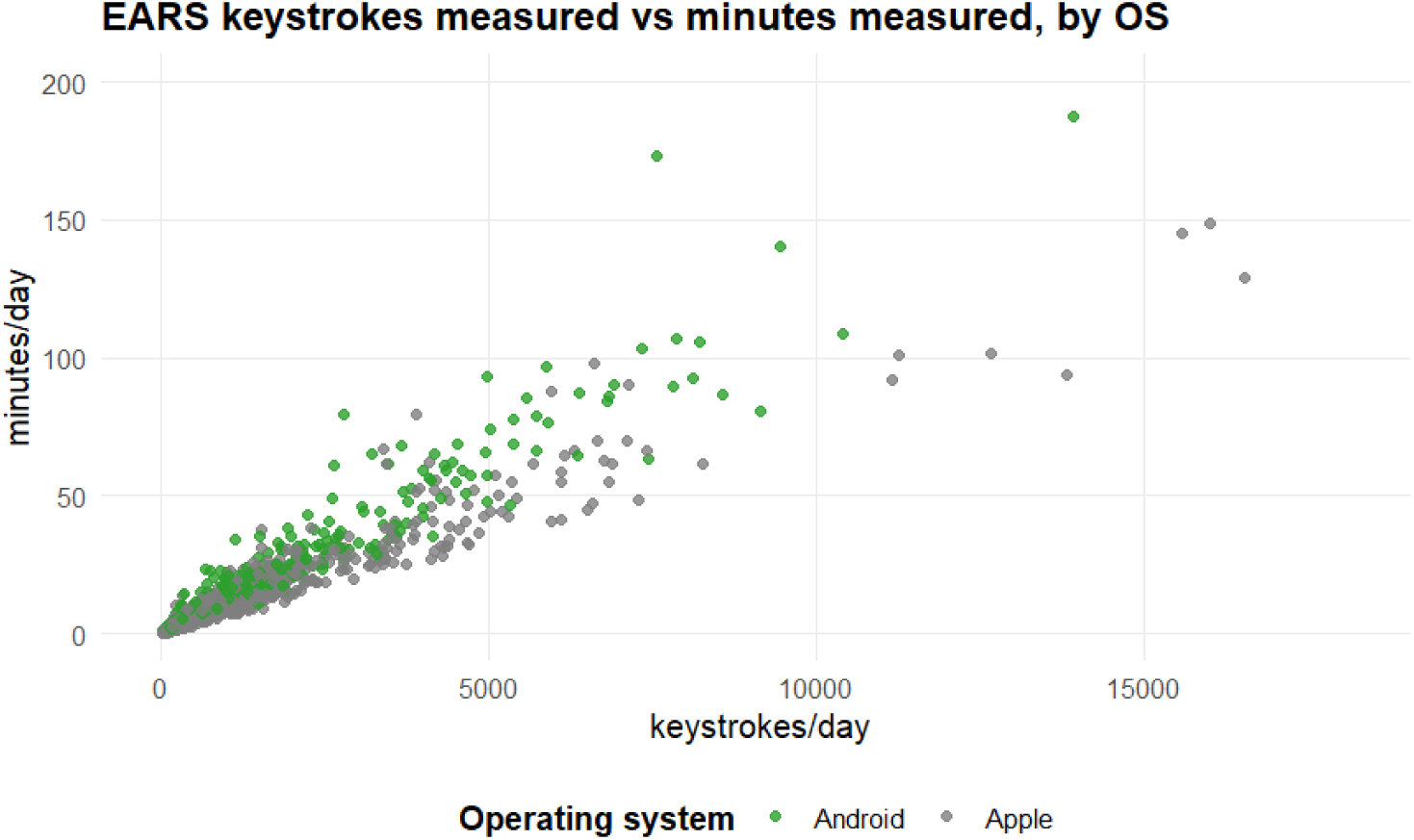
Teens who use iPhones have been recorded with lower average keystrokes and total minutes of smartphone usage compared to Android users.

**Figure 6.**
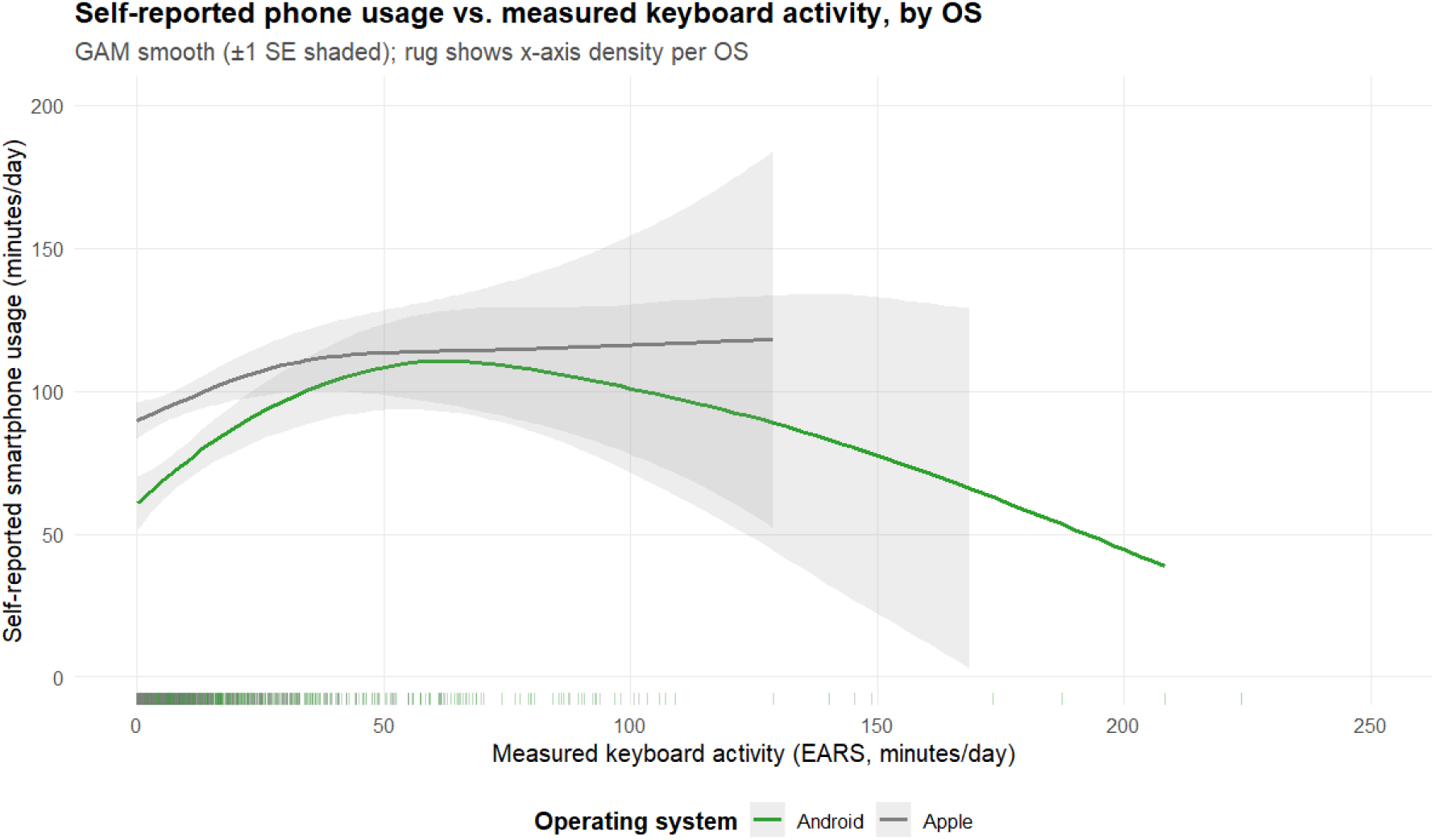
displays the relationship between EARS-captured number of keyboard usage minutes and self-reported minutes of smartphone usage (smoothed), for both operating systems. At the lower end of usage (< 50 min), iPhone users (gray) self-reported higher smartphone usage than Android users (green).

Given our sample size constraints, we chose to analyze both operating systems together and included OS as a control variable in the subsequent analysis. While iOS keystrokes may underestimate true smartphone use, they still capture meaningful behavior patterns: higher keystrokes correlate with higher self-reported use for both operating systems, and systematic differences between OS diminish at higher usage levels.

### Temporal Considerations (Protocol vs. Non-protocol)

Beyond determining which participants to include, we also needed to establish the appropriate temporal window for data inclusion. Participants received Fitbit devices before the protocol period began and retained them for a period afterward, resulting in varying amounts of out-of-protocol wear data across participants. We compared data collected during protocol versus non-protocol periods to determine whether to use all Fitbit observations or prioritize data quality by restricting the data to protocol periods only.

Figures Figure 7, Figure 8, and Figure 9 show that participants exhibited similar behaviors within-protocol period (within dashed bars) but differed drastically from the before/post-protocol observations. We concluded that we could not assume data from the within-protocol and the out-of-protocol wears come from the same distribution and decided to only include the Fitbit observations from the protocol periods. At the conclusion of this preprocessing stage, we resolved to use within-protocol Fitbit activity and sleep data with at least 5 weekdays and 2 weekend days based on our principled approach to person-level thresholding and treating outliers.

**Figure 7.**
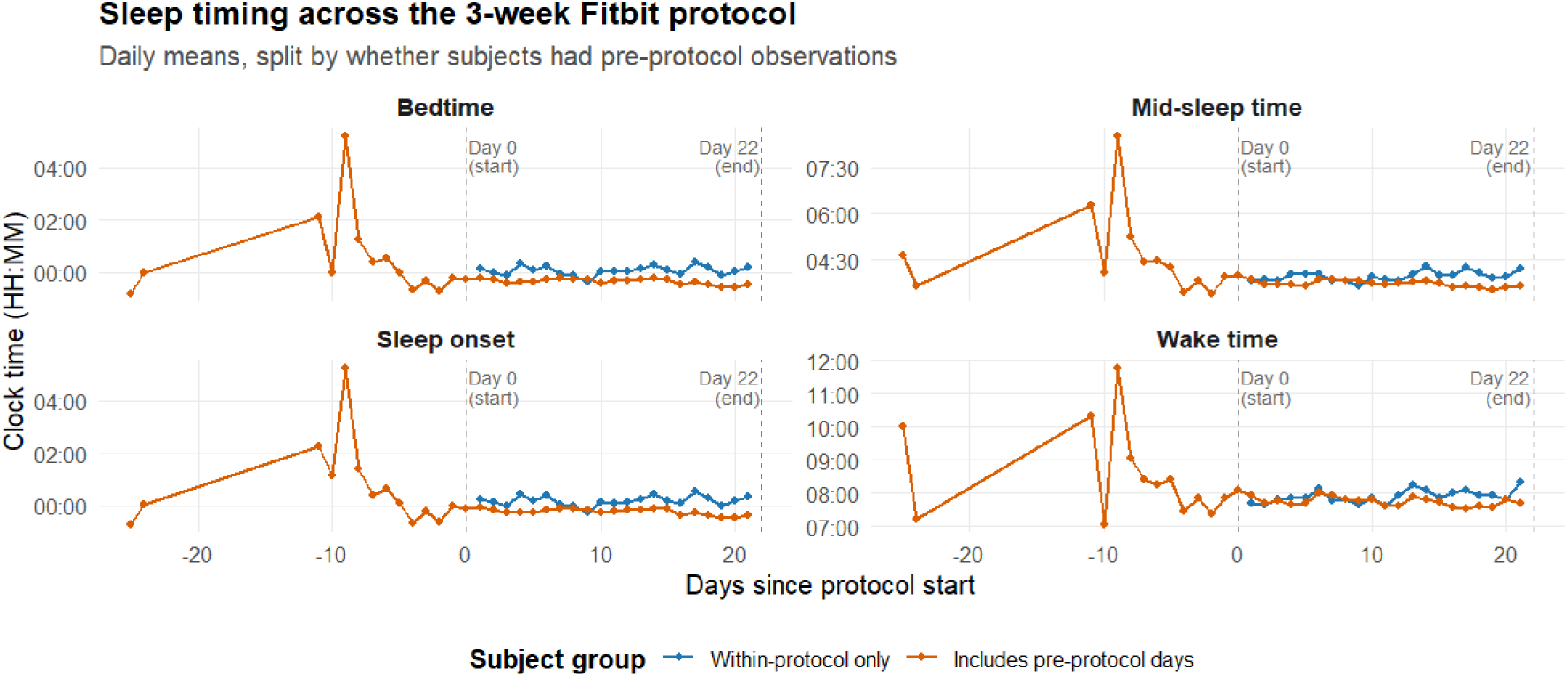
In the Fitbit sleep dataset, we compared the day-level observations of participants with before-protocol wear (blue) and that of participants who only had within-protocol wear data (orange). All participants exhibited similar behaviors within the protocol period (within dashed bars) but differed drastically from the before protocol observations. Sleep data has been pre-cleaned by the ABCD team to exclude post-protocol wear.

**Figure 8.**
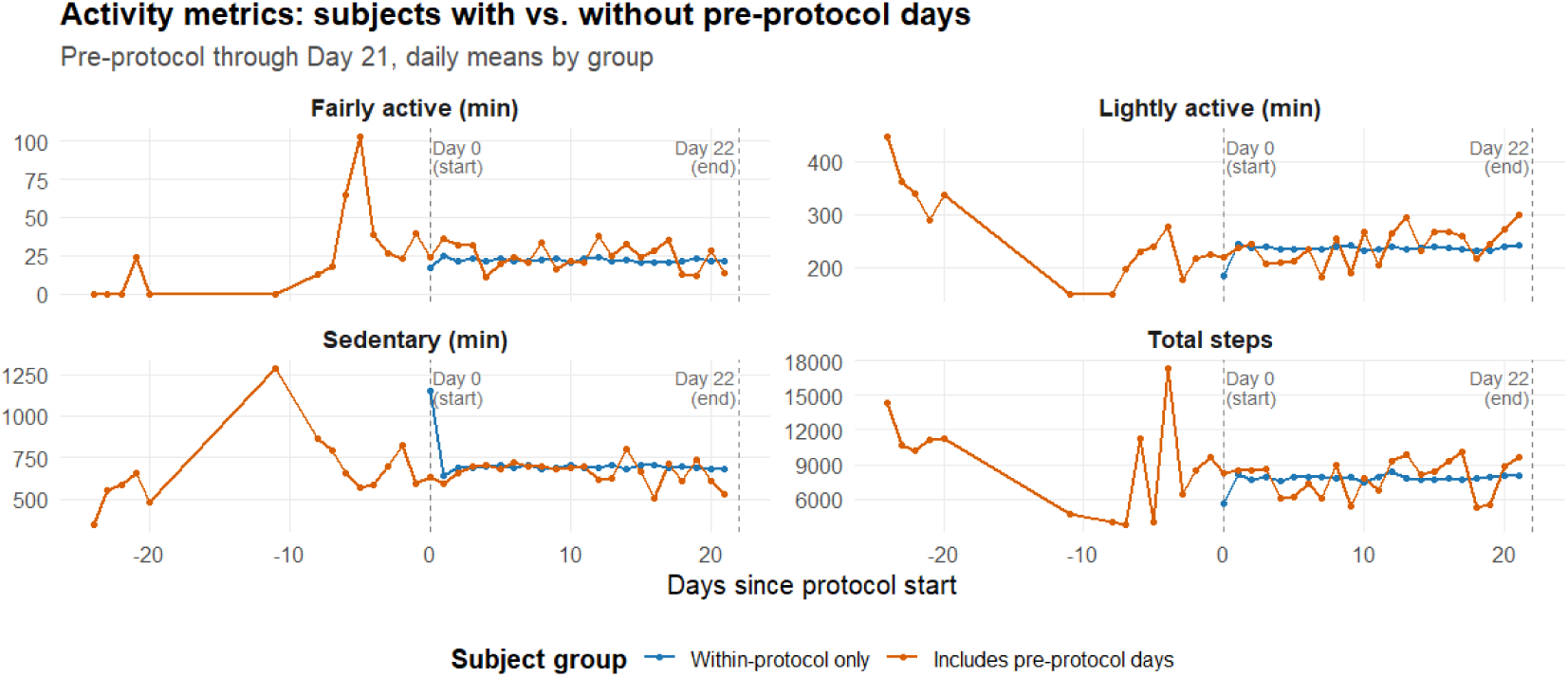
compared the day-level observations of participants with before-protocol wear (orange) and that of participants without before-protocol wear data (blue). All participants exhibited similar behaviors within the protocol period (within dashed bars) but differed drastically from the before/after protocol observations.

**Figure 9.**
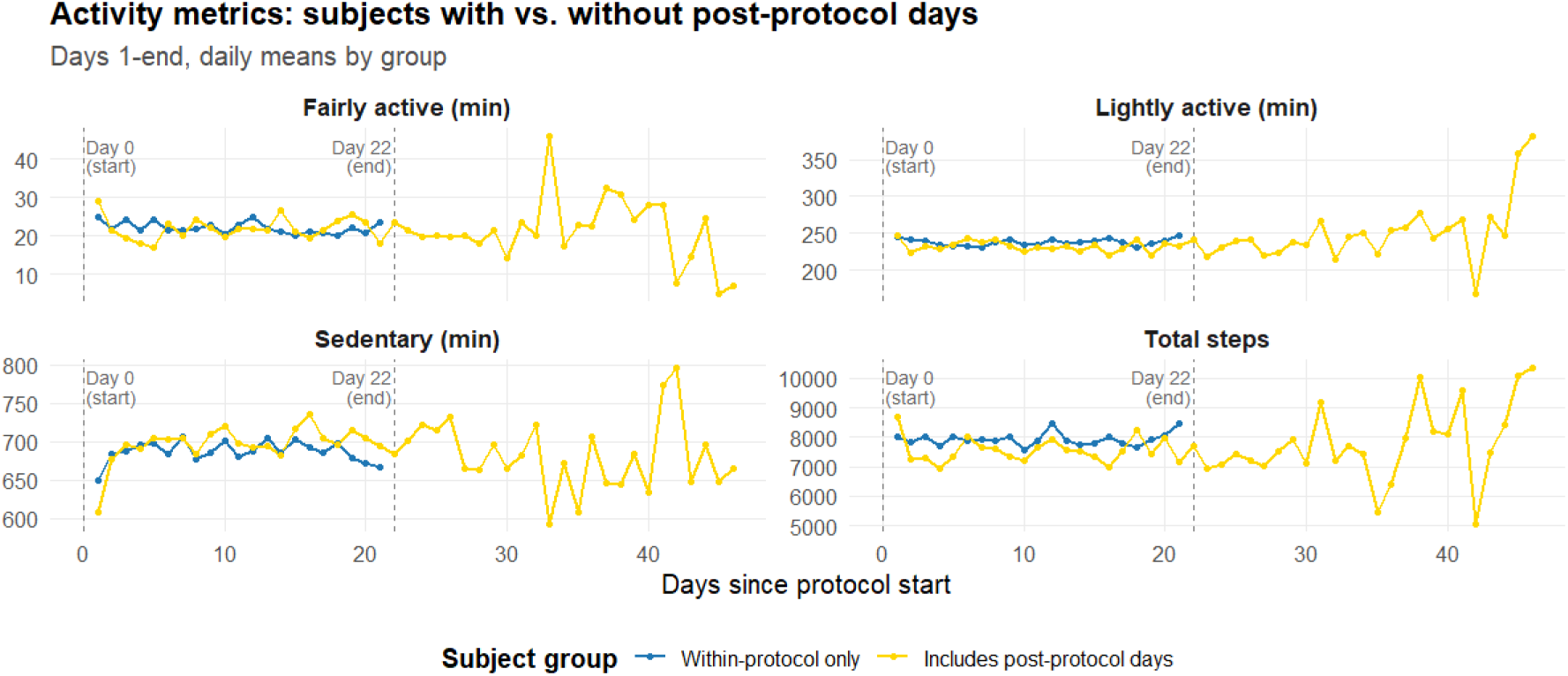
compared the day-level observations of participants with post-protocol wear (yellow) and that of participants without pre-protocol wear data (blue). All participants exhibited similar behaviors within the protocol period (within dashed bars) but differed drastically from the before/after protocol observations

For EARS data on smartphone use, the summary data that is more readily accessible to researchers are summarized from hourly raw data. Unfortunately, in contrast to the Fitbit sleep and physical activity data, we found that the provided EARS dataset summarizes all observed days during the usage tracking period (which encompasses but extends beyond the protocol period) with at least one upload, offering no method to isolate protocol-specific data. Figure 10 shows that many data points are comprised of more than 21 days of observations, indicating that non-protocol days are included in the summary measures. Data collected during protocol periods were monitored by ABCD researchers [19] and therefore likely represent higher data quality than data collected during non-protocol periods. Based on our analyses with the Fitbit data, it may be the case that only within-protocol data should be used. However, processing these data is time intensive and highly burdensome (especially in the case of App Usage data which involves extensive preprocessing by the ABCD team) and doing so likely goes beyond the scope of many research projects. Although outside the scope of this project, we recommend that future research attend to this issue. At minimum, it would be helpful to researchers to mention the extent of out-of-protocol data included in the summary data to enhance data transparency and guide future projects leveraging these data, rather than excluding a discussion of out-of-protocol data as has been the case in prior ABCD EARS reports[19].

**Figure 10.**
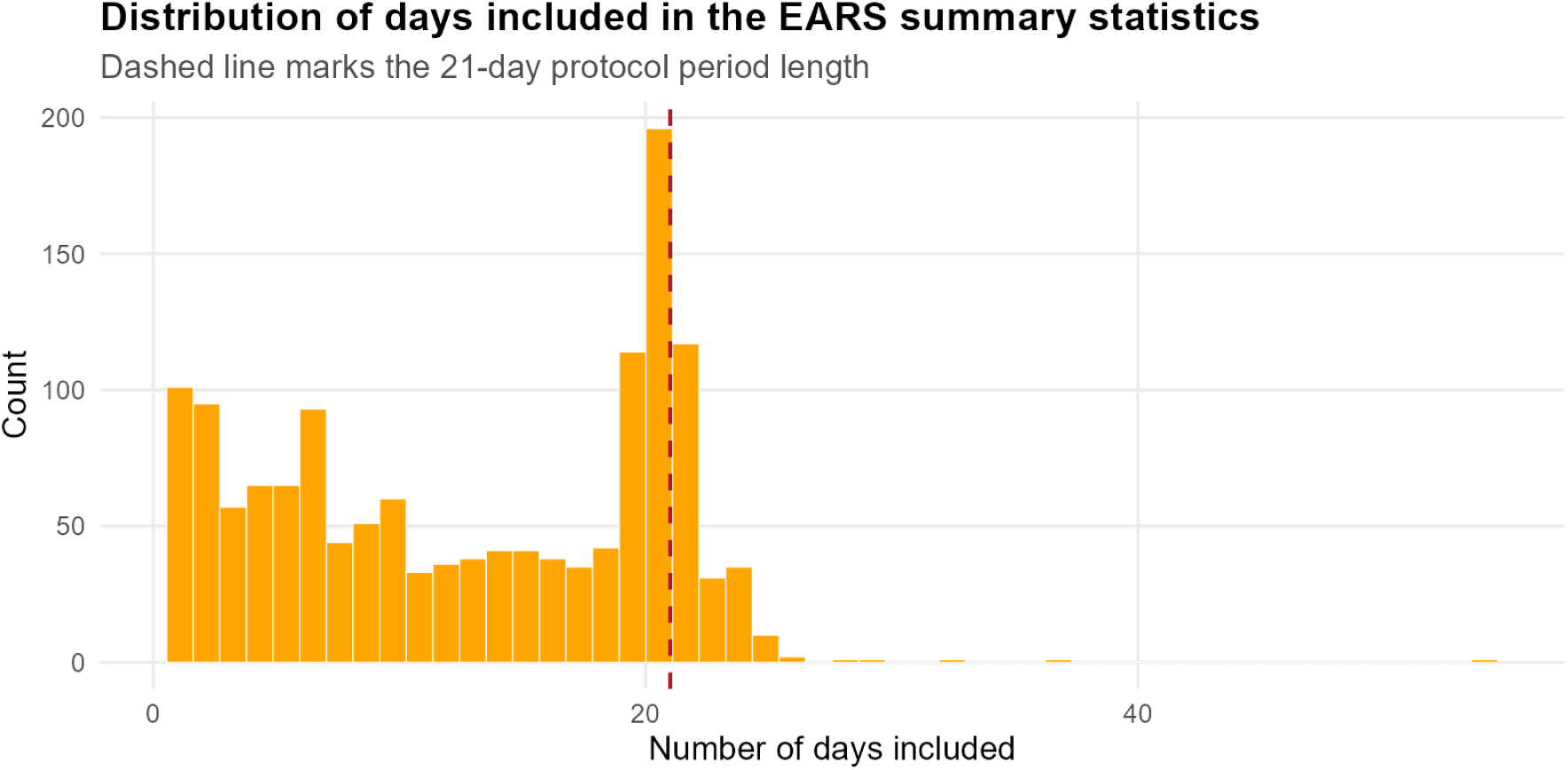
Distribution of the number of days included in the EARS summary measures. Bars right of the dashed line indicate nontrivial number of data points containing observations outside of the protocol window.

### Feature Engineering and Variable Selection

Our next task was to select specific variables for our machine learning model. For many machine learning tasks, such as clustering, it is beneficial to maximally capture all potentially relevant features from the candidate variables. Research using passively sensed data and machine learning commonly considers a wide array of derived features for each variable of interest [33]. For example, one could calculate summary statistics (e.g., mean, median, *SD*, min, max) across the observation period for each individual to capture nuanced characteristics of the underlying distributions, each reflecting distinct aspects of behavior that may hold predictive value. To illustrate, the intraindividual spread of sleep time (i.e., consistency of a teen’s bedtime each night) provides different information than the mean (i.e., a teen’s average sleep time). Prior research [34, 35] suggesting that weekend and weekday sleep and activity levels represent distinct constructs, particularly in adolescents; therefore, we created features for weekends and weekdays separately. For the Fitbit sleep and physical activity data, we calculated the mean, median, standard deviation, range, minimum, and maximum across observed days for weekend and weekday periods separately for each participant. We also included additional sleep variables following a prior study [36]: sleep onset latency (time to fall asleep), sleep inertia (time to get out of bed after first waking), sleep efficiency (ratio of total time asleep to total time in bed), and social jetlag (difference between average mid-sleep point on weekends versus weekdays).

#### Treatment of sleep timing variables

Defining various summary statistics for sleep timing variables (e.g., sleep time, wake up time) is challenging because time wraps around midnight [37]. For example, 11:59 pm is only two minutes from 12:01 am, but common representation of time such as in statistical programs (which tracks time from a reference point, usually midnight) would incorrectly calculate the distance as 23 hours and 58 minutes, making calculations of means, standard deviations, or simple differences in the Euclidean space meaningless. To address this issue, the ABCD team offset all sleep-timing variables by 12 hours so that midnight is represented as noon (i.e., 00:00 = noon; 23:59 = 11:59 AM). However, this simply moves the cusp from midnight to noon is not robust against high-spread samples [37]. In the ABCD dataset, we observe adolescents who sleep past noon or stay awake all night and go to sleep in the morning hours, creating outliers that cause the same issue.

Instead, we converted time data to polar coordinates (representing time as positions on a 24-hour circular scale) using the R package *circular* [37–39], performed the desired calculations to derive time differences, and converted back to the time scale. However, defining *max* and *min* sleep times remained problematic: if someone went to sleep at noon one day, is that their minimum (considering it a very early bedtime) or maximum (if they stayed awake all night, making it very late)? Moreover, we needed to define distances in a mixture of angular (time) and Euclidean data for clustering.

To address these challenges, we calculated deviations from reference sleep (10:00 pm) and wake times (7:00 am) as surrogates for the time data based on typical sleep schedules for adolescents. While ideal sleep times vary based on chronotypes and school schedules, these reference points provide a consistent baseline across all participants, and the variability in deviations still informs us about sleep timing consistency. The choice for these reference sleep and wake times may certainly differ depending on a researcher’s specific sample, setting, and aims.

#### Selection of EARS app categories

We calculated the percentage of people with 0 usage (in all three measures *key, min, ses*) in each of the app categories (Figure 11). To reduce the number of covariates, we only included categories with at least 10% non-zero keystroke observations, thus excluding art, business, finance, food, medical, sports, and weather. These rarely-used categories aligned with the perceptions of adolescents on a youth advisory board with whom we consulted during this project, thus validating our decision to exclude these categories.

**Figure 11.**
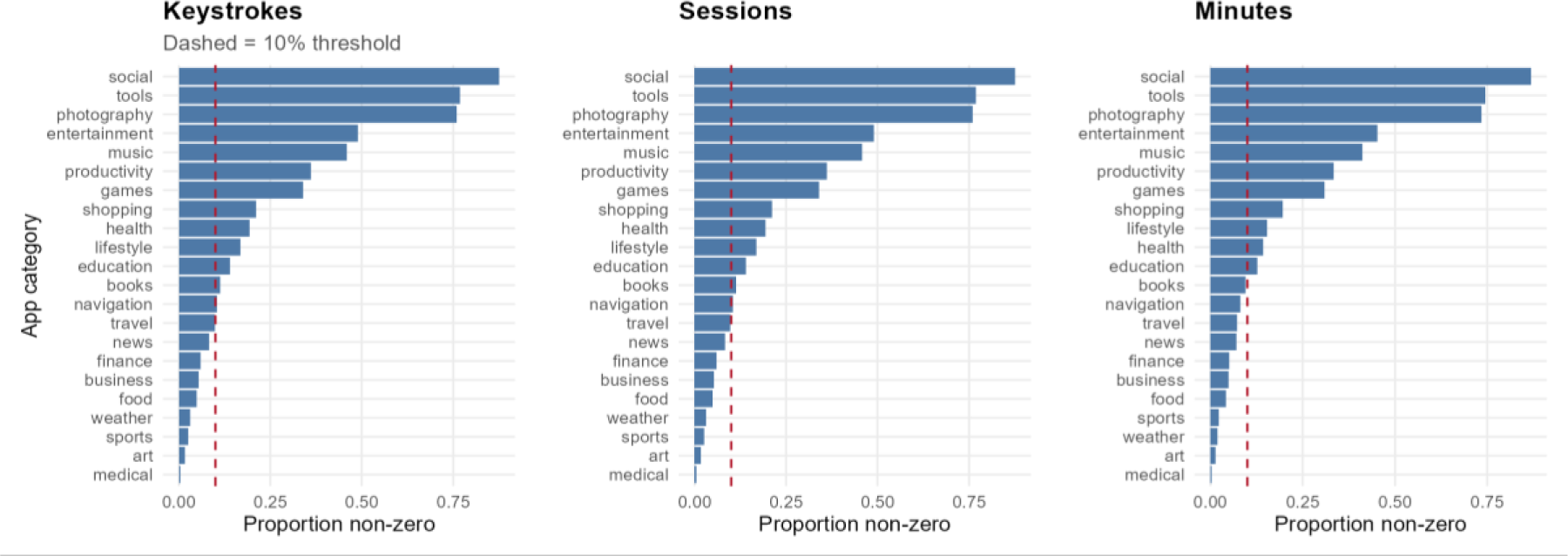
Percentage of participants with no measured usage in each of the app categories calculated separately for keyboard measures: Keystrokes, Sessions, and Minutes.

#### Proportion of participants with non-zero app usage by category

### Other Quality Control Challenges

Any project seeking to leverage vast amounts of passive data will encounter additional quality control challenges and decision points. For this project, in the Fitbit dataset’s week-level summaries, we noted contradictory data encoding. For heart rate-based activity minutes and total steps, two similar measurements exist: *per day* (Fitbit built-in) versus *avg* (ABCD team calculation). The *avg* variables had more NA values than *per day* variables, and these NAs were incorrectly coded as 0s during week-level aggregation (Figure 12). Inadvertently treating missing values as 0 could have biased overall findings. To ensure proper data control, we used day-level data directly and selected Fitbit *per day* variables over ABCD *avg* measures.

**Figure 12.**
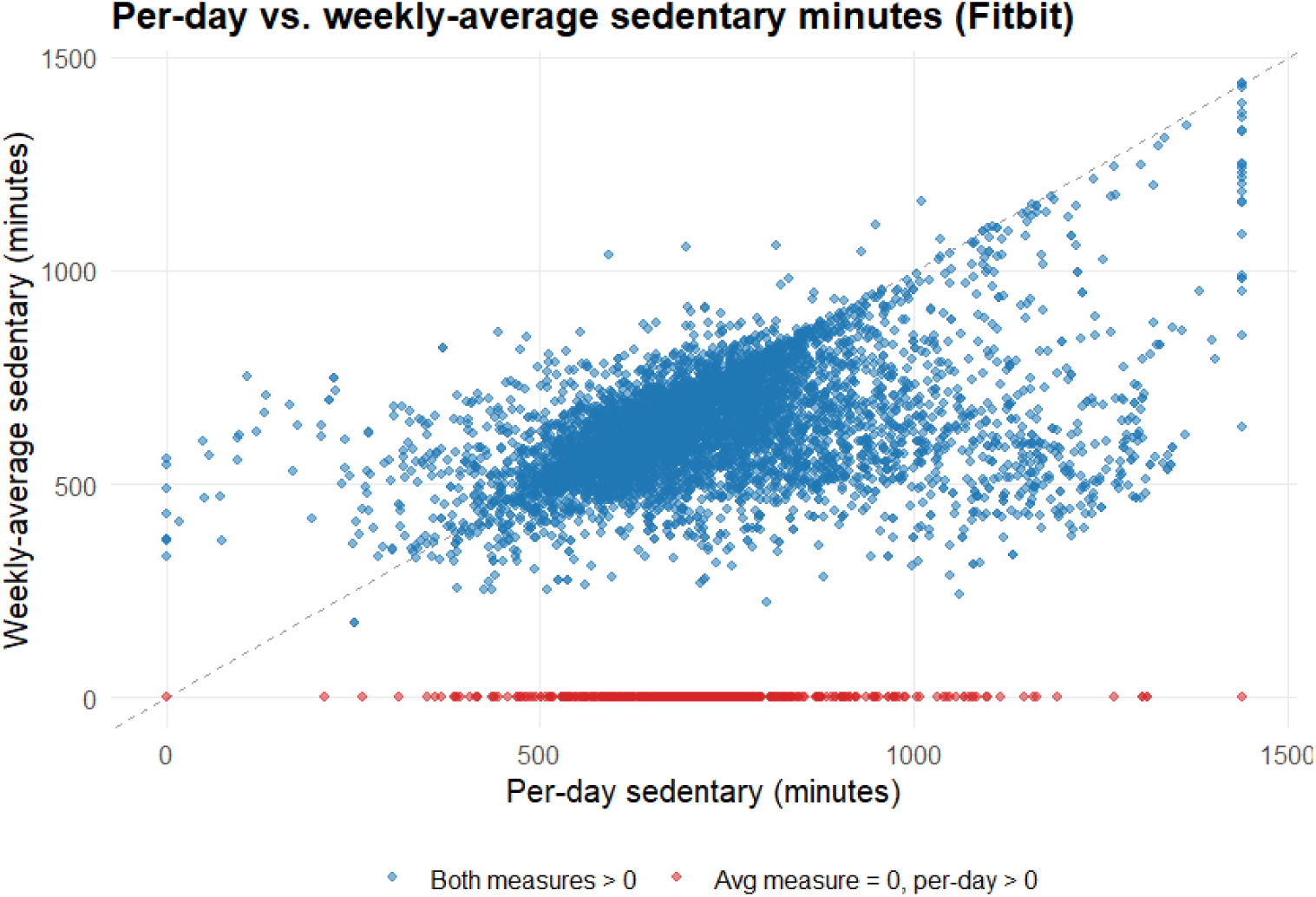
is an example of missing values incorrectly coded as 0 (red) in the week level summary for avg sedentary min (Y-axis) when the observation is not missing in per-day sedentary minutes (X-axis).

Additionally, in the day-level summary datasets, we found individuals with “duplicated” sleep entries on the same day of week 1 but with different bedtimes and wear dates, which contradicts the ABCD documentation. Investigation into the protocol start date and the wear date revealed that the week number was assigned incorrectly. We recoded the week numbers based on the days since the start of the protocol for each individual (i.e., the first 7 days are coded as week 1). While time-consuming, this careful combing through the data was necessary to catch errors and inconsistencies that were not captured in the ABCD documentation.

### Post-Processing Data

All EARS and Fitbit measures were aggregated to the participant level (N = 428) with a final set of features (number of features = 380) distinguishing weekends vs. weekdays. The average age of the final sample of participants was 14.1 years (SD: 0.7), 55% were female, and the race/ethnicity breakdown is as follows: participants identified as: White: 271 (63.3%), Non-white Hispanic: 78 (18.2%), Black: 29 (6.8%), Asian: 2 (0.4%), Other: 48 (11.2%). Table 3 displays the demographic composition of the samples before and after preprocessing. We note that 8 participants were excluded due to missing genetic data to create polygenic risk scores. This exclusion is relevant to our analysis aim but is omitted from the main text given our primary focus on passively sensed data.

**Table 3.**
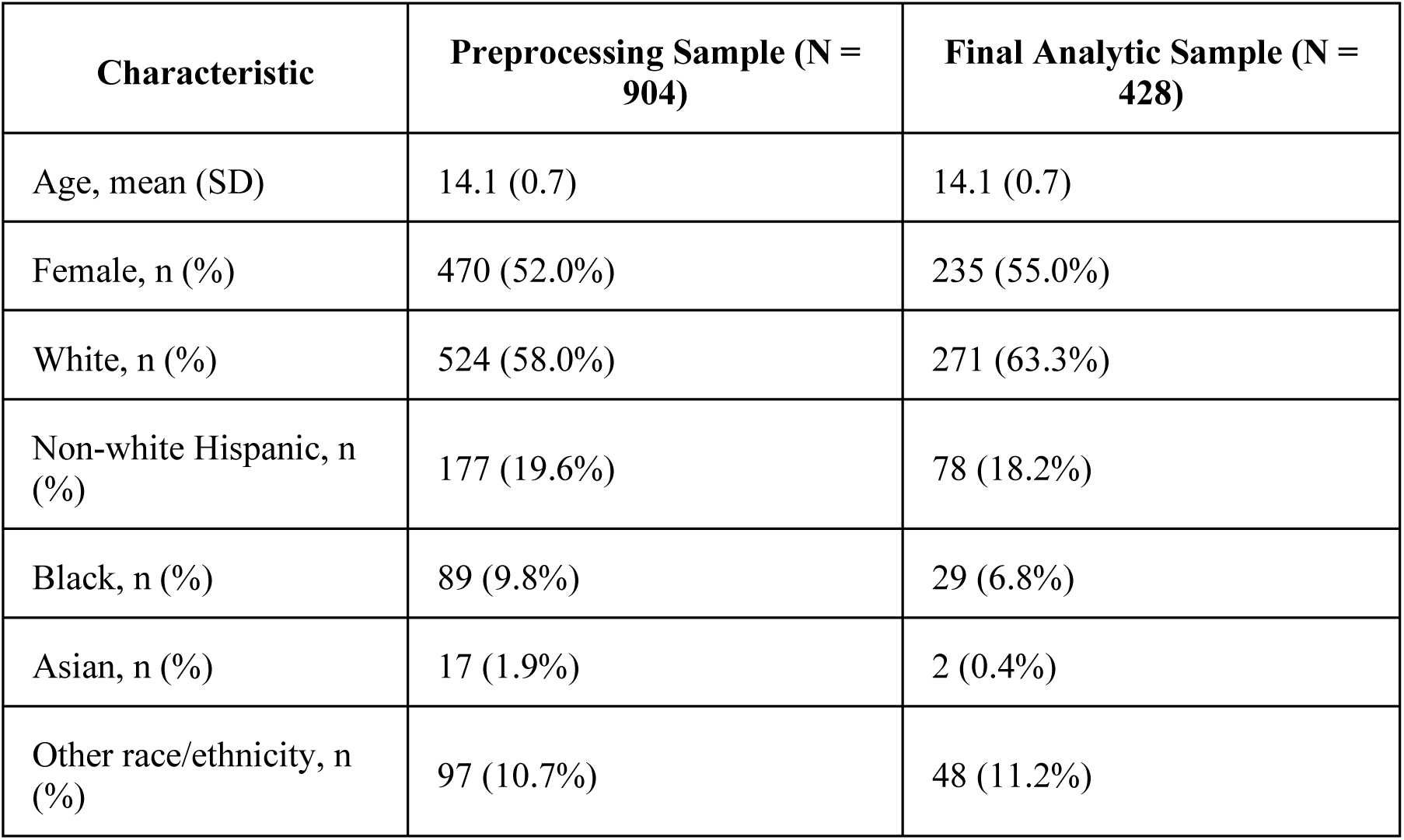
Demographic Characteristics of the Preprocessing and Final Analytic Samples.

| Characteristic | Preprocessing Sample (N = 904) | Final Analytic Sample (N = 428) |
| --- | --- | --- |
| Age, mean (SD) | 14.1 (0.7) | 14.1 (0.7) |
| Female, n (%) | 470 (52.0%) | 235 (55.0%) |
| White, n (%) | 524 (58.0%) | 271 (63.3%) |
| Non-white Hispanic, n (%) | 177 (19.6%) | 78 (18.2%) |
| Black, n (%) | 89 (9.8%) | 29 (6.8%) |
| Asian, n (%) | 17 (1.9%) | 2 (0.4%) |
| Other race/ethnicity, n (%) | 97 (10.7%) | 48 (11.2%) |

#### Demographic Characteristics of the Preprocessing and Final Analytic Samples

## Discussion

The decisions made during data cleaning and feature engineering can substantially influence analytic results, yet these steps are often relegated to supplementary materials or omitted entirely from published articles. By providing a detailed account of our preprocessing approach, we hope to contribute to methodological transparency in digital phenotyping research.

Our framework offers three key recommendations for researchers working with passive sensing data. First, favor person-based inclusion criteria over rigid variable-based cutoffs. Variable-based thresholds risk discarding extreme observations that reflect genuine behavioral variability rather than measurement error. In our data, extreme values were concentrated among participants with sparse observations, suggesting that engagement can be informative for exclusion. Second, examine sample distributions over time rather than assuming observations are exchangeable. Data collected outside the protocol window are common, but evidence is limited on whether such out-of-protocol data should be included.

Although more data may seem preferable, our Fitbit analyses showed systematic differences between observations collected within versus outside the protocol window, leading us to exclude out-of-protocol Fitbit data. Future studies should carefully evaluate this issue to determine which observations to retain in final analyses. Third, thoroughly investigate and report data quality issues. Our review of the ABCD pipeline surfaced errors not noted in the ABCD documentation, including missing values coded as zeros in week-level aggregates and incorrectly assigned week numbers. These issues would have biased downstream results had they gone undetected.

While illustrated through the ABCD Study, these principles apply broadly to research leveraging smartphones, wearables, and other digital data sources. We hope this work serves as both a practical guide for ABCD data users and a methodological contribution to passive sensing data preprocessing.

## Limitations

Several limitations should be noted. First, our minimum observation threshold (5 weekdays, 2 weekend days) was informed by prior literature and domain expertise but remains somewhat arbitrary; different thresholds would yield different samples and might be more appropriate for different contexts. Moreover, our person-based cut-offs drastically reduced sample sizes, while this is a substantial trade-off, we believe a smaller but more reliable dataset is sufficient for our pilot study given that the other half of the year 4 follow up data will become available in future releases. Furthermore, the person-based approach assumes that Fitbit engagement patterns are unrelated to our outcomes of interests. If adolescents who wear their Fitbit less consistently are at higher risk for substance use initiation, this approach may systematically exclude high-risk individuals, potentially biasing our findings. Second, restricting to protocol periods only for the Fitbit datasets created discrepancies between Fitbit and the smartphone data since they were not aggregated in the same fashion; however, we considered that they are inherently different measurement tools aimed at capturing personal traits. Third, we were unable to fully resolve the source of operating system differences in EARS data, leaving potential bias unaddressed. Finally, while our approach is designed to be generalizable, specific decisions (e.g., reference sleep times, app category harmonization) may require adaptation for different populations, devices, or research aims.

## Conclusion

Passively sensed data from wearable devices and smartphones hold substantial promise for advancing substance use prevention and intervention research. However, researchers are often confronted with equally substantial challenges in understanding, processing, and cleaning these complex data streams. As illustrated throughout this paper, even in large-scale studies with extensive documentation, researchers must make non-trivial decisions regarding data inclusion, quality, and valid observation windows. These decisions should be informed by both the characteristics of the target population (e.g., adolescents) and the intended downstream analyses (e.g., clustering for substance use initiation prediction). We hope this work provides practical guidance for researchers working with similar forms of passive sensing data and encourages greater transparency and documentation of preprocessing and analytic workflows in digital phenotyping research.

## Acknowledgement

The authors thank Dr. Brant P. Hasler for his guidance on the minimum number of days of observation required to reliably estimate social jetlag, which informed our participant-level inclusion criteria. We also thank the members of the Youth Research Advisory Board for their input throughout this work, which helped ground our data decisions in youth perspectives and ensure the relevance of the derived features.

## Statements and declarations

### Ethical considerations

This research was approved by the Institutional Review Board at the University of Pittsburgh (STUDY24050100).

### Consent to participate

N/A.

### Consent for publication

N/A.

### Data availability

Data used in the preparation of this article were obtained from the Adolescent Brain Cognitive DevelopmentSM (ABCD) Study (https://abcdstudy.org), held in the NIMH Data Archive (NDA). This is a multisite, longitudinal study designed to recruit more than 10,000 children age 9-10 and follow them over 10 years into early adulthood. The ABCD Study® is supported by the National Institutes of Health and additional federal partners under award numbers U01DA041048, U01DA050989, U01DA051016, U01DA041022, U01DA051018, U01DA051037, U01DA050987, U01DA041174, U01DA041106, U01DA041117, U01DA041028, U01DA041134, U01DA050988, U01DA051039, U01DA041156, U01DA041025, U01DA041120, U01DA051038, U01DA041148, U01DA041093, U01DA041089, U24DA041123, U24DA041147. A full list of supporters is available at https://abcdstudy.org/federal-partners.html. A listing of participating sites and a complete listing of the study investigators can be found at https://abcdstudy.org/consortium_members/. ABCD consortium investigators designed and implemented the study and/or provided data but did not necessarily participate in the analysis or writing of this report. This manuscript reflects the views of the authors and may not reflect the opinions or views of the NIH or ABCD consortium investigators. The ABCD data repository grows and changes over time. The ABCD data used in this report came from DOI 10.15154/z563-zd24. DOIs can be found at https://nda.nih.gov/abcd/abcd-annual-releases.

## References

[1] Insel TR. Digital Phenotyping: Technology for a New Science of Behavior. JAMA 2017; 318: 1215–1216.

[2] Cornet VP, Holden RJ. Systematic review of smartphone-based passive sensing for health and wellbeing. J Biomed Inform 2018; 77: 120–132.

[3] Bagot KS, Matthews SA, Mason M, et al. Current, future and potential use of mobile and wearable technologies and social media data in the ABCD study to increase understanding of contributors to child health. Dev Cogn Neurosci 2018; 32: 121–129.

[4] Behrendt S, Wittchen H-U, Höfler M, et al. Transitions from first substance use to substance use disorders in adolescence: Is early onset associated with a rapid escalation? Drug Alcohol Depend 2009; 99: 68–78.

[5] Maggs JL, Staff J, Patrick ME, et al. Very early drinking: Event history models predicting alcohol use initiation from age 4 to 11 years. Addictive Behaviors 2019; 89: 121–127.

[6] Ystrom E, Kendler KS, Reichborn-Kjennerud T. Early age of alcohol initiation is not the cause of alcohol use disorders in adulthood, but is a major indicator of genetic risk. A population-based twin study. Addiction 2014; 109: 1824–1832.

[7] Shen SY, Qi W, Zeng J, et al. Passive Sensing for Mental Health Monitoring Using Machine Learning With Wearables and Smartphones: Scoping Review. J Med Internet Res 2025;27:e77066 https://www.jmir.org/2025/1/e77066 2025; 27: e77066.

[8] Baroudi L, Zernicke RF, Tewari M, et al. Using Wear Time for the Analysis of Consumer-Grade Wearables’ Data: Case Study Using Fitbit Data. JMIR Mhealth Uhealth; 13. Epub ahead of print 2025. DOI: 10.2196/46149.

[9] Yang FN, Picchioni D, Duyn JH. Effects of sleep-corrected social jetlag on measures of mental health, cognitive ability, and brain functional connectivity in early adolescence. Sleep; 46. Epub ahead of print 1 December 2023. DOI: 10.1093/SLEEP/ZSAD259.

[10] Zhang R, Schwandt ML, Vines L, et al. Changes in Rest-Activity Rhythms in Adolescents as They Age: Associations With Brain and Behavioral Changes in the ABCD Study. J Am Acad Child Adolesc Psychiatry 2025; 64: 1056–1067.

[11] Castiglione-Fontanellaz CEG, Schaufler S, Wild S, et al. Sleep regularity in healthy adolescents: Associations with sleep duration, sleep quality, and mental health. J Sleep Res 2023; 32: e13865.

[12] West AB, Bittel KM, Russell MA, et al. A systematic review of physical activity, sedentary behavior, and substance use in adolescents and emerging adults. Transl Behav Med 2020; 10: 1155–1167.

[13] Hasler BP, Smith LJ, Cousins JC, et al. Circadian rhythms, sleep, and substance abuse. Sleep Med Rev 2012; 16: 67–81.

[14] Halabi R, Selvarajan R, Lin Z, et al. Comparative Assessment of Multimodal Sensor Data Quality Collected Using Android and iOS Smartphones in Real-World Settings. Sensors 2024; 24: 6246.

[15] Slade C, Sun Y, Chao WC, et al. Current challenges and opportunities in active and passive data collection for mobile health sensing: a scoping review. JAMIA Open 2025; 8: ooaf025.

[16] Trifan A, Oliveira M, Oliveira JL. Passive Sensing of Health Outcomes Through Smartphones: Systematic Review of Current Solutions and Possible Limitations. JMIR Mhealth Uhealth 2019; 7: e12649.

[17] Lind MN, Kahn LE, Crowley R, et al. Reintroducing the Effortless Assessment Research System (EARS). JMIR Ment Health 2023;10:e38920 https://mental.jmir.org/2023/1/e38920 2023; 10: e38920.

[18] Lind MN, Byrne ML, Wicks G, et al. The Effortless Assessment of Risk States (EARS) Tool: An Interpersonal Approach to Mobile Sensing. JMIR Ment Health 2018;5(3):e10334 https://mental.jmir.org/2018/3/e10334 2018; 5: e10334.

[19] Alexander JD, Linkersdörfer J, Toda-Thorne K, et al. Passively sensing smartphone use in teens with rates of use by sex and across operating systems. Sci Rep; 14. Epub ahead of print 1 December 2024. DOI: 10.1038/S41598-024-68467-8.

[20] Wade NE, Ortigara JM, Sullivan RM, et al. Passive Sensing of Preteens’ Smartphone Use: An Adolescent Brain Cognitive Development (ABCD) Cohort Substudy. JMIR Ment Health; 8. Epub ahead of print 1 October 2021. DOI: 10.2196/29426.

[21] Kim N, Li LY, Funkhouser CJ, et al. EEG-based brain connectivity and sentiment analysis from smartphone social communication: insights into remitted major depressive disorder among adolescents. NPP - digital psychiatry and neuroscience; 3. Epub ahead of print 7 October 2025. DOI: 10.1038/S44277-025-00044-X.

[22] Ng MY, Frederick JA, Fisher AJ, et al. Identifying Person-Specific Drivers of Depression in Adolescents: Protocol for a Smartphone-Based Ecological Momentary Assessment and Passive Sensing Study. JMIR Res Protoc 2024; 13: e43931.

[23] Zhang Y, Pratap A, Folarin AA, et al. Long-term participant retention and engagement patterns in an app and wearable-based multinational remote digital depression study. NPJ Digit Med 2023; 6: 25.

[24] Lee J, Turchioe MR, Creber RM, et al. Phenotypes of engagement with mobile health technology for heart rhythm monitoring. JAMIA Open 2021; 4: ooab043–ooab043.

[25] Garavan H, Bartsch H, Conway K, et al. Recruiting the ABCD sample: Design considerations and procedures. Dev Cogn Neurosci 2018; 32: 16–22.

[26] Haist F, Jernigan TL. Adolescent Brain Cognitive Development Study (ABCD) - Annual Release 5.1, 10.15154/z563-zd24 (2024, accessed 25 May 2026).

[27] Godino JG, Wing D, de Zambotti M, et al. Performance of a commercial multi-sensor wearable (Fitbit Charge HR) in measuring physical activity and sleep in healthy children. PLoS One 2020; 15: e0237719.

[28] Novel Technologies, https://web.archive.org/web/20240918152055/https://wiki.abcdstudy.org/release-notes/non-imaging/novel-technologies.html#fitbit-physical-activity-daily (accessed 13 December 2025).

[29] Consortium A. ABCD Data Documentation. DOI: 10.5281/ZENODO.17154440.

[30] Wing D, Godino JG, Baker FC, et al. Recommendations for Identifying Valid Wear for Consumer-Level Wrist-Worn Activity Trackers and Acceptability of Extended Device Deployment in Children. Sensors (Basel); 22. Epub ahead of print 1 December 2022. DOI: 10.3390/S22239189.

[31] Zhang L, Oshri A, Carvalho C, et al. Prospective associations between sleep, sensation-seeking, and mature screen usage in early adolescents: findings from the Adolescent Brain Cognitive Development study. Sleep; 48. Epub ahead of print 1 January 2025. DOI: 10.1093/SLEEP/ZSAE234.

[32] Fischer D, Klerman EB, Phillips AJK. Measuring sleep regularity: theoretical properties and practical usage of existing metrics. Sleep; 44. Epub ahead of print 11 October 2021. DOI: 10.1093/SLEEP/ZSAB103.

[33] Sameh A, Rostami M, Oussalah M, et al. Digital phenotypes and digital biomarkers for health and diseases: a systematic review of machine learning approaches utilizing passive non-invasive signals collected via wearable devices and smartphones. Artificial Intelligence Review 2024 58:2 2024; 58: 66-.

[34] Sun W, Ling J, Zhu X, et al. Associations of weekday-to-weekend sleep differences with academic performance and health-related outcomes in school-age children and youths. Sleep Med Rev 2019; 46: 27–53.

[35] Treuth MS, Catellier DJ, Schmitz KH, et al. Weekend and Weekday Patterns of Physical Activity in Overweight and Normal-weight Adolescent Girls. Obesity (Silver Spring) 2007; 15: 1782.

[36] Kiss O, Baker FC, Palovics R, et al. Using explainable machine learning and fitbit data to investigate predictors of adolescent obesity. Sci Rep; 14. Epub ahead of print 1 December 2024. DOI: 10.1038/S41598-024-60811-2.

[37] Klerman EB, Wang W, Phillips AJK, et al. Statistics for Sleep and Biological Rhythms Research: Longitudinal Analysis of Biological Rhythms Data. J Biol Rhythms 2016; 32: 18.

[38] Jammalamadaka SR, SenGupta A. Topics in Circular Statistics. 5. Epub ahead of print April 2001. DOI: 10.1142/4031.

[39] Lund U, Agostinelli C. Circular Statistics [R package circular version 0.5-2]. CRAN: Contributed Packages. Epub ahead of print 25 September 2025. DOI: 10.32614/CRAN.PACKAGE.CIRCULAR.

